# Impaired visual processing in psychosis patients with a predisposition for visual hallucinations

**DOI:** 10.1101/2022.05.05.22274713

**Authors:** Marouska van Ommen, Jan Bernard Marsman, Remco Renken, Richard Bruggeman, Teus van Laar, Frans W. Cornelissen

## Abstract

Psychosis is frequently associated with the occurrence of visual hallucinations (VH), but their etiology remains largely unknown. While patients with psychosis show deficits on various behavioral visual and attentional tasks, previous studies have not specifically related these deficits to the presence of VH. This suggests that tasks used in these studies do not target the visual-cognitive neural mechanisms that mediate VH, which in turn limits the development of effective therapies. We therefore designed a study to target these mechanisms directly. In this case control study we asked patients with psychosis who had previously experienced VH to indicate when they recognized objects that were gradually emerging from dynamic visual noise, while scanning their brains using functional Magnetic Resonance Imaging. In a previous study, this recognition task was used to identify the neural basis of VH in patients with Parkinson’s Disease. Based on this earlier work, we decided to test the following hypothesis: when compared to psychosis patients not experiencing VH and age-matched healthy controls, psychosis patients with VH show reduced occipital activity and frontal activity around the moment of recognition (known as pop-out). For all groups, neuroimaging revealed increased activity in all examined visual areas around pop-out. However, psychosis patients with VH showed reduced occipital responsiveness, especially in the inferior part of the bilateral lateral occipital complex, a region known to play a key role in object recognition. We did not observe altered frontal or prefrontal activity before pop-out in this group. A possible explanation is that the relatively sustained activation of the visual memory-related angular gyri around pop-out may have compensated for the impaired early visual processing in psychosis patients with VH. We discuss our results in terms of current theories of visual hallucinations, such as predictive coding and contextual modulation. Our study is the first to show that visual processing deficits contribute to the occurrence of VH in psychosis. These findings could be used to develop tests to identify the visual-cognitive mechanisms that mediate VH in this group.

## Introduction

Patients with schizophrenia have chronic or recurrent psychoses ^1^. Hallucinations are a core symptom and are sensory experiences while being awake, without corresponding external stimulation of the relevant sensory organ ^2^. Most are auditory in nature, with visual hallucinations (VH) being the second most common type. They have a lifetime prevalence of 37.1% ^3^. VH in psychosis are mostly complex, involving animals and people ^4,5^. They can be frightening ^5^. Their underlying cognitive and neural mechanisms remain largely unknown. Insight into these in patients with psychosis with a predisposition for VH may increase our understanding of the cause of their hallucinations and help to improve therapy.

In general, schizophrenia is associated with impairments in various aspects of visual processing related to object perception, visual attention and working memory. Compared to healthy controls, patients with schizophrenia have reduced surround suppression in early visual areas during a contrast-contrast illusion ^6^, are worse at detecting contours embedded in noise ^7^, visually integrating a contour consisting of individual elements ^8,9^ and recognizing faces ^10^, colors and novel shapes ^11,12^. This suggests that patients with schizophrenia have impaired object perception. Moreover, they are worse at guiding attention to task-relevant inputs ^13^, at indicating a certain letter combination in a stream of random letters ^10^, and at indicating which of many squares changed colour ^14^. This suggests that patients with schizophrenia have impaired attention and working memory function. However, it is unknown whether and how such general impairments relate to the occurrence of VH. Our own previous research using behavioral testing and addressing facial affect recognition, sustained attention and attentional switching, found no differences between patients with and without VH ^3^. The only difference was that patients without VH had worse sustained attention scores compared to controls, whereas VH patients did not. Overall, these behavioral tasks results provided no evidence for impaired visual (or early visual) or attentional processing that is specific to patients experiencing VH. An issue may be that current clinical tests may lack the sensitivity to detect subtle differences that could identify causal mechanisms.

One way to overcome this and to increase our understanding of both common and differential mechanisms underlying VH is to assess different patient populations using neuro-imaging in combination with the same tasks ^3,15^. Previously, our group studied VH in Parkinson’s Disease (PD). We found that VH in PD were related to behavioral impaired visual object and space perception, and impaired sustained attention ^16,17^. To identify possible neural correlates, we combined an object recognition task, the ‘Image Recognition Movies’ (IRM), with fMRI ^18^. In this task, a visual object gradually appears out of dynamic visual noise. The task was designed such as to extend the uncertainty associated with, and the processing underlying, visual perception. This temporal extension made it possible to acquire informative data with the low temporal resolution of fMRI. Moreover, our assumption was that differences in identified neural mechanisms during normal object recognition would provide us with pointers towards potentially abnormal processing causal to VH. In the task, participants are asked to identify the moment they recognize the object. Prior to recognition, HC, VH patients and non-VH patients showed increased activity in visual areas. However, compared to HC and non-VH patients, before recognition, VH patients had a smaller increase in activity in the lateral occipital cortex (LOC), ventral temporal cortex, inferior parietal cortex, right superior frontal gyrus and middle frontal gyrus. This suggests that in PD, the occurrence of VH relates to both impaired visual and top-down processing.

In the present study we assessed visual perception and attention in patients with psychosis using the same tasks described above for patients with PD. We also assessed working memory. Based on our previous clinical testing, we first tested two hypotheses: 1) compared to HC, psychosis patients have worse behavioral performance, and 2) compared to each other, psychosis patients with a predisposition for VH and patients without VH have the same behavioral performance. Based on our previous PD findings and using the same fMRI experimental design, we subsequently tested a third hypothesis: 3) prior to image recognition, patients with a predisposition for VH – when compared to those without VH – show reduced responsiveness of the occipital areas, including the object recognition region LOC, and reduced activation of the frontal areas.

## Materials and methods

The study consisted of two parts. In part 1, participants performed behavioural tests. In part 2, participants performed an fMRI experiment. Approximately two-third of the participants who participated in part 2 also participated in part 1. In the results section, we therefore separately report the characteristics and behavioral scores of these two groups.

### Participants

This study belongs to the INZICHT study, which goal is to gain insight into VH in psychosis. It has a behavioral part (INZICHT1, https://www.trialregister.nl/trial/4858), and an fMRI part (INZICHT2, https://www.trialregister.nl/trial/6685). The ethics board of the University Medical Center Groningen (UMCG) approved the study protocol. All participants provided written informed consent. The study followed the tenets of the Declaration of Helsinki. Participants received a 25 euro coupon for participating in INZICHT1, and a 50 euro coupon for INZICHT2.

Participants were recruited via: the GROUP study ^19^, the Department of Psychotic Disorders UMCG, Lentis Center for Mental Health Groningen and Winschoten, and Anoiksis, a patients association of psychotic disorders. Healthy controls (HC) were also recruited via word of mouth advertising. Participants for INZICHT1 were asked to participate in INZICHT2. Participants fulfilled the following criteria: age between 18-55; speaking Dutch fluently, being able to give informed consent. In addition, patients met DSM-IV-TR criteria (or the DSM-5 equivalent) for schizophrenia, schizophreniform disorder, schizoaffective disorder or psychotic disorder not otherwise specified (NOS) ^20^. For INZICHT1: patients had VH more than a couple of times in the last month (belonging to the patient group with a predisposition for VH, PSVH+), never had VH (PSVH−), or had VH but did not meet the PSVH+ criteria (PSVH-prone). Because of the relatively low signal-to-noise ratio in fMRI, INZICHT2 did not include the intermediate PSVH-prone category. Due to difficulties in recruiting patients, inclusion criteria for the PSVH− group were somewhat less strict than INZICHT1: two PSVH− participants in INZICHT2 had experienced VH: the first once around 7 years ago, the other participant twice 8 years ago. In case of psychiatric comorbidity, the psychotic disorder had to be predominating. Moreover, the VH had to be psychosis related. Their own psychiatrist evaluated these last two conditions. Exclusion criteria were psychiatric and neurological disorders that presumably affect the data, visual acuity <0.5 (tested with a chart with sentences presented at reading distance), visual field defects (Donders’ confrontational technique, cognitive impairment (Mini-Mental State Examination score <26 ^21^, for INZICHT2: fMRI incompatibility. Furthermore, HC were excluded if they or a first degree family member who has or had psychosis, or if they themselves ever had VH. The groups were age, gender and cognition matched (MMSE>25). For both part 1 and 2 visual acuity was corrected if necessary.

### Procedure

#### Part 1: Behavioral tests

##### Visual perception

The validated Visual Object and Space Perception Battery (VOSP) assessed object and space perception ^22,23^. Object perception included Incomplete Letters, Silhouettes, Object Decision and Progressive Silhouettes; space perception comprised Dot Counting, Position Discrimination, Number Localisation and Cube Analysis. Per subtest, the number of correct answers was determined. The Progressive Silhouettes test assessed the number of figures necessary for recognition.

##### Attention

The subtest ‘Vigilance’ (Test battery of Attentional Performance (TAP)), measured sustained visual attention ^24^. During this 10-minute computer task, a pattern alternately popped up in the upper and lower square. Participants pushed a button for irregularities. The reaction time and the number of omitted irregularities were measured.

##### Working memory

The Groninger Word Learning Task (WLT) assessed working memory ^25,26^. Fifteen words were presented on a computer screen. Afterwards, participants were asked thrice to recall all words; and again after 20 minutes (delayed recall). Then, 30 words were verbally presented (spoken). Participants indicated whether these words were new or not (delayed recognition).

##### Auditory perception

As patients with VH frequently also have auditory hallucinations (11), auditory perception was assessed as well, using The Speech Discrimination Task (SDT) ^27^. For 100 trials, first target words, superimposed on white background noise, were presented. Then, a second word was presented, without noise. Participants were asked whether the words were the same; 1 meant ‘certainly not’, 3 ‘I don’t know’, 5 ‘certainly’. The number of hits, false alarms and ‘don’t know’s’ were measured. The mean certainty of answers (range 3-5) was extracted.

#### Image Recognition Movies task (IRM)

The Image Recognition Movies task (IRM) included 4 × 15 movies of 30 seconds, during which a static image of an animal or object gradually appeared out of dynamic random visual noise ^16^. The moment of image recognition, ‘pop-out’, was indicated by pushing a button on the response box with the right index finger. Participants verbally named the image. The speed and content of recognition (hit, false alarm, don’t know) were assessed. During the presentation of the movies, a central fixation square changed color at random intervals. Participants were asked to push another button as quickly as possible following such a color change, to keep attention constant. Moreover, their response time on this task was subtracted from the recorded pop-out time, to derive recognition time. In between movies, a red fixation cross on a dark background was presented. The task was practiced before testing until participants understood the task correctly (no-one needed more than two practice movies). If <80% of HC recognized a movie, it was excluded (this applied to 10 movies). Four participants, three PSVH+ and one PSVH− participant, were excluded because of repetitive (>30 times) pushing for recognition. One participant was excluded because the content of the answer was not assessed. Moreover, movies were excluded for which participants pushed more than once, if there was a mismatch between behavioral data and logfiles (for example having named the image, but not pushing the button, or vice versa), or technical problems. The mean included movies for PSVH+ was 48.1, for PSVHprone 47.4, for PSVH− 47.3, and for HC 48.2.

#### Part 2: fMRI

##### Behavioral and ophthalmic data

Participants were interviewed about psychotic symptoms using the Questionnaire for Psychotic Experiences (QPE; http://qpeinterview.com/home/ ^28^) and the validated Positive and Negative Syndrome Scale (PANSS, ^29^) in Dutch. Moreover, different aspects of their vision were assessed (if necessary, vision was corrected): visual acuity (digital Snellen chart), contrast sensitivity (GECKO chart, maximum score 16) and visual fields (Humphrey Field Analyzer, 24-2 SITA; Carl Zeiss Meditec).

##### fMRI and MRI acquisition

The IRM was presented during fMRI, using a different set of 50 movies (22 animals, 22 objects and six meaningless objects (control)). Movies were projected on a translucent screen, which participants viewed via a mirror. Stimuli covered approximately 18° of the visual field. Movies were presented in a fixed order in 2 runs of 25 movies each. In contrast to the behavioral variant, in this fMRI variant of the IRM, participants did press a button on recognition, but did not name the image. Thus, the content of recognition, ‘correct’ or ‘wrong’ was not assessed. See Fig. 1A for a schematic depiction of the paradigm. fMRI data were collected using a 3T Philips Magnetic Resonance system with a standard 64-channel SENSE head coil (Intera, Philips Medical Systems, Best, The Netherlands). Echo-planar images were acquired: anterior-posterior, 39 slices per volume, TR 2 s, TE 30 ms, voxel size 3 mm isotropic, flip angle 90**°** (FOV 192×192×117 mm). Furthermore, an anatomical T1 was acquired (160 slices, voxel size 1 mm isotropic, FOV 256×224×160 mm). After scanning, participants were interviewed about the occurrence of any hallucinations during scanning.

**Fig. 1:**
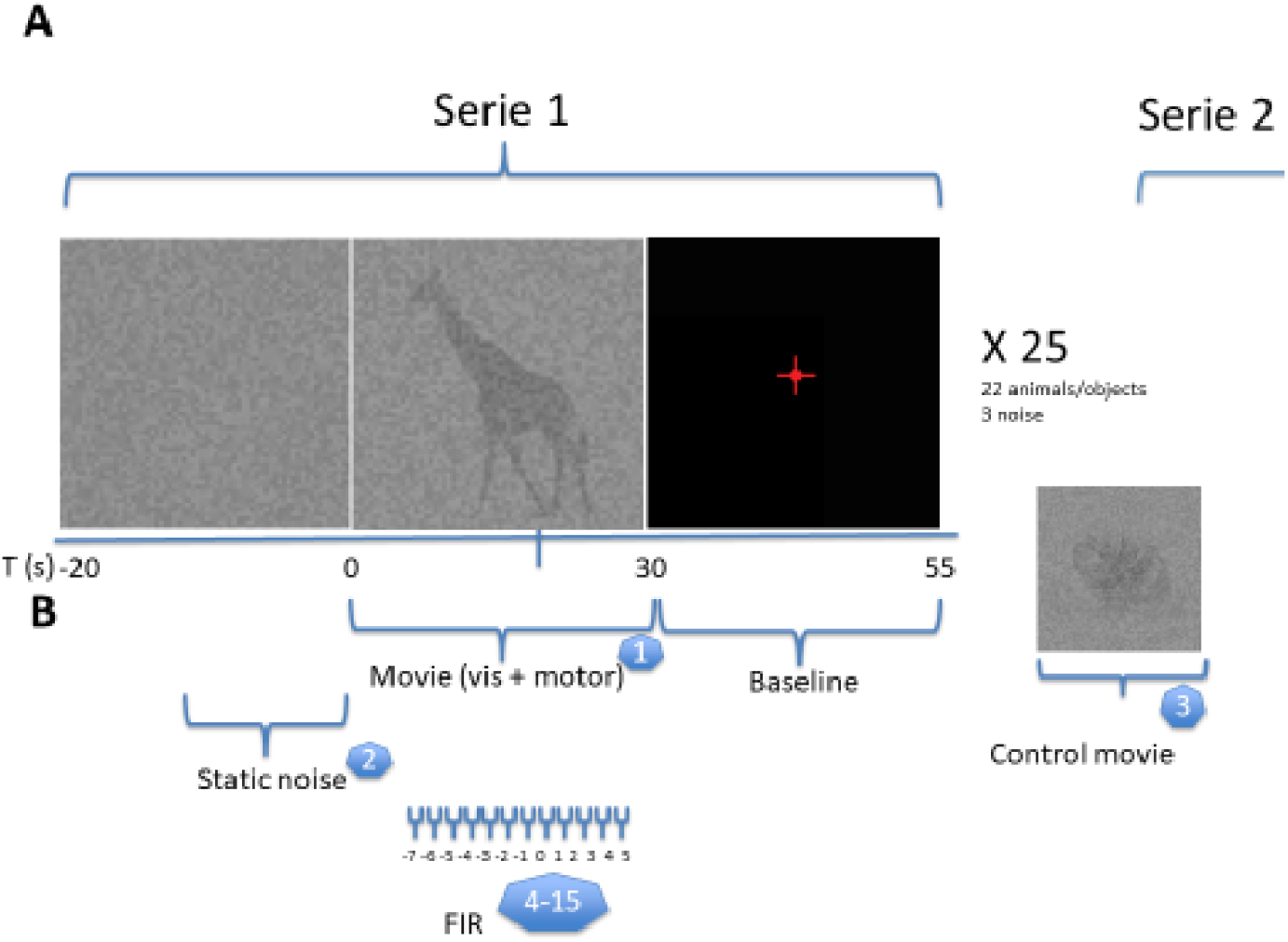
Image Recognition Movies task: A) paradigm, B) model for FIR analyses. Legend: <u>A</u>) The Image Recognition Task paradigm ^18^. A screen, viewed via a mirror, displayed 50 movies during which a static image (22 animals, 22 well-known objects and six meaningless objects (control)) gradually pops out of random noise. The movies covered approximately 18° of the visual field. The task included 2 runs of 25 movies; each movie lasted 30 seconds. In between movies, a red fixation cross on a dark background was presented. Image recognition, ‘pop-out’, was indicated by a button push. During the movies a central fixation square changed color with random intervals. Participants pushed another button for this color change, to keep attention constant. <u>B</u>) Finite Impulse Response (FIR) analyses determined the temporal dynamics of visual recognition ^18^. These include mean whole brain activations per time-bin around pop-out. These were generated per group from 7 seconds before pop-out until 5 seconds after pop-out (12 time-bins of each 1 second (=0.5 TR)). Besides these bins (being the 4^th^-15^th^ condition), 3 other conditions were modeled: 1) movies, 2) control movies, 3) static noise at the beginning of the runs. ‘Movies’ was included as a baseline to ensure that the FIR analyses addressed pop-out related activity instead of overall task-effects or general neuronal excitability.

#### Statistical analyses

Analyses of behavioral and clinical data were performed with SPSS v20 ^30^. Demographic and illness characteristics were compared using Fisher’s Exact Tests and Pearson Chi-Square tests for categorical variables, and the non-parametric Mann-Whitney U test and Kruskal-Wallis tests for continuous variables. Test score comparisons are presented by MANOVAs (Wilks’ lambda), although they were not normally distributed. Non-parametric tests showed similar results, but do not reflect relationships between dependent variables. If both multivariate and subsequent univariate testing were significant, non-parametric tests were used to further explore differences between the groups, using Bonferroni correction (part 1: SD/4, part 2: SD/3). We also compared test scores without the PSVH-prone group, as this group is small and may therefore confound the results. As VH in psychosis frequently co-occur with auditory hallucinations ^4^, Spearman correlations determined the correlation between hits on the IRM and SDT. All analyses were two-tailed with α=0.05.

##### fMRI and MRI pre-processing and analyses

The anatomical scans were co-registered, normalized to Montreal Neurological Institute (MNI) space, and segmented. Functional scans were realigned, co-registered to the MNI-152 EPI template, and normalized using the deformation field obtained from the segmentation. Subsequently, spatial smoothing (8 mm) and denoising (ICA-AROMA ^31^) was applied. Data were analyzed using SPM12 (Wellcome centre human neuroimaging), running in MATLAB 2014b ^32^. All fMRI analyses were time-locked to the moment of pop-out (the recorded pop-out time corrected for the mean reaction time on the square changing colour). In case participants had pressed the button to indicate recognition more than once, the time of the first button press was taken. The main analysis aimed at investigating the activity patterns around pop-out. The additional analysis, that focussed on the effect of recognition (i.e. examining differences in activity pattern from pop-out until movie end), is reported on in Suppl. 1.

Finite Impulse Response (FIR) analyses determined whole brain activity at a temporal resolution of 1 sec (=1/2 TR) around pop-out (Fig. 1B). These 1-sec time-bins were generated per group from 7 sec before pop-out until 5 sec after pop-out (for a total of 12 time-bins). General Linear Models (GLMs) were built per participant, with 15 conditions in the following order: 1) movie, 2) control movie, 3) static noise at the beginning of each run, 4-15) the time-bins. The condition ‘movie’ was included as a baseline to ensure that the FIR addressed pop-out-related activity instead of overall task-effects or general neuronal excitability. Per subject, the contrast ‘movie’ versus baseline (fixation cross) was obtained. Next, group means were determined (Suppl. 2A1,B1). Subsequently, ANOVA compared groups for ‘movie’ with p<0.001 on the voxel level and 0.05 FWE cluster-wise correction (Suppl. 2A2,B2).

Last, activity patterns during these time-bins were statistically explored for predefined Regions Of Interest (ROIs) that are involved in visual processing. As the task includes recognizing complex images, we included both hierarchically early and later visual areas. ROIs were based on the Harvard-Oxford atlas; only the LGN ROI was based on the Juelich histological atlas ^33^. ROI time series were plotted per group. Next, mean group activity was compared between groups per time-bin, applying the following contrasts: PSVH+ versus PSVH−, PSVH+ versus HC, PSVH− versus HC. ROI analyses were two-tailed with p<0.05 and corrected for the number of time-bins and contrasts using FWE correction.

### Data, stimulus and code availability

Data sharing is available upon request and follows UMCG guidelines. IRM movies can be obtained via www.visualneuroscience.nl. Code is available on request.

## Results

To preview our main results, in the behavioural testing performed of part 1, we found that in general, patients performed worse than the HC-group. However, no differences were found between the three different patient groups (PSVH+, PSVHprone and PSVH−). Despite the absence of any marked behavioural differences in the PSVH+ group, in the fMRI experiment of part 2, we found evidence for a reduced responsiveness of the occipital visual processing regions, most markedly in the LOC region involved in object recognition. Moreover, PSVH+ showed a tendency for reduced suppression of the angular gyri around image recognition. Below, we describe these results in detail.

### Part 1: Behavioral tests

Table 1A lists the demographic and illness characteristics of all participants. Sixty-five participants were included: 18 PSVH+, 10 PSVH-prone, 17 PSVH− and 20 HC. Of the patients, 32 had schizophrenia, 9 schizoaffective disorder, 3 psychotic disorder NOS and 1 schizophreniform disorder. The total patient group had a mean age of 35.9 years (SD 9.5); HC 33.9 (SD 10.9). Of all patients 66.7% was male, compared to 70% of the HC. Patients had a lower mean education level than HC (6.7 (SD 1.3) versus 7.4 (SD 0.8), p<.05). The mean disease duration was 13 years (SD 19.2, range 1-34), without significant differences between the patient groups (p=.23). Most patients used antipsychotics, with the majority using 1 atypical antipsychotic.

**Table 1:**
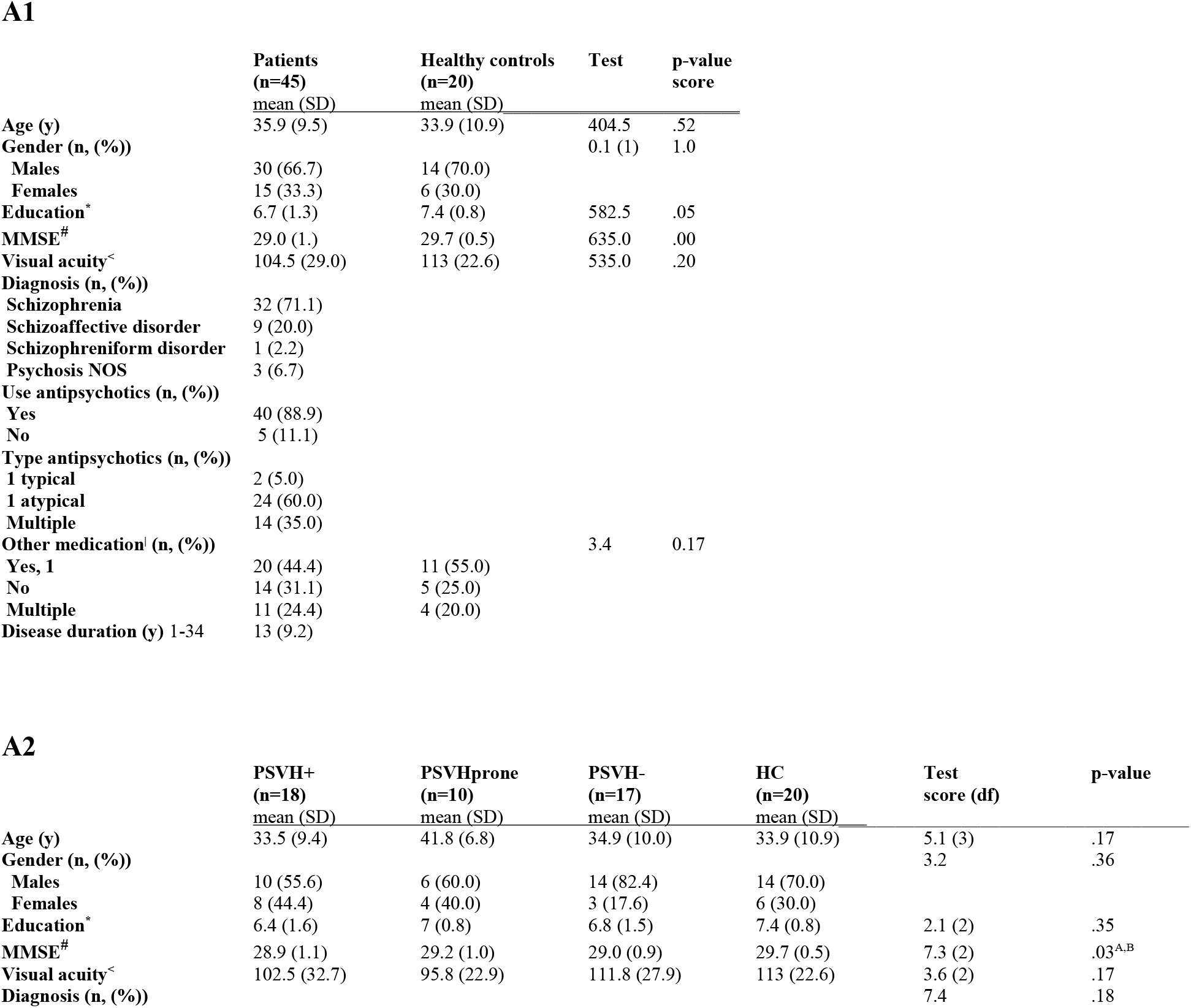
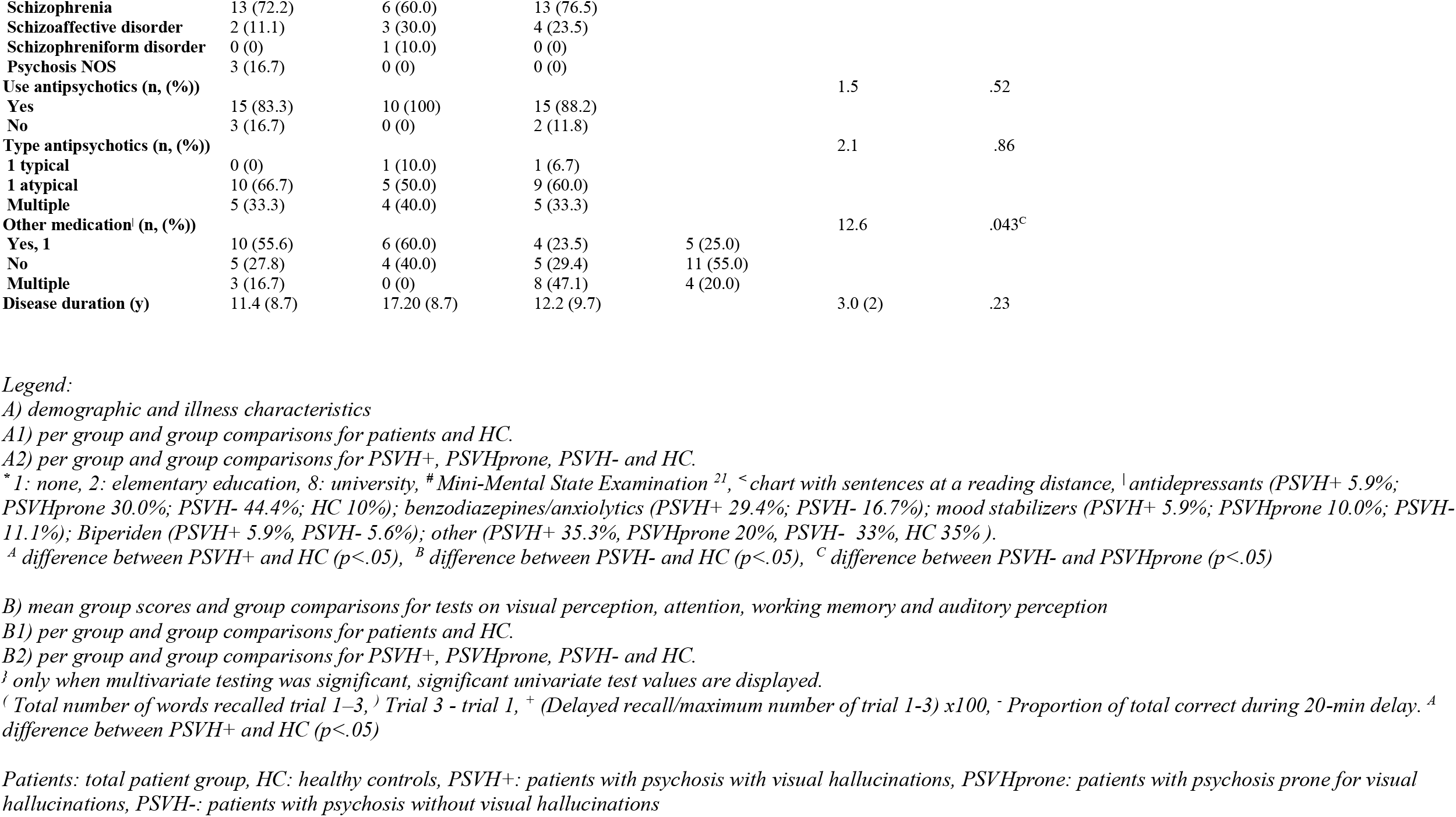
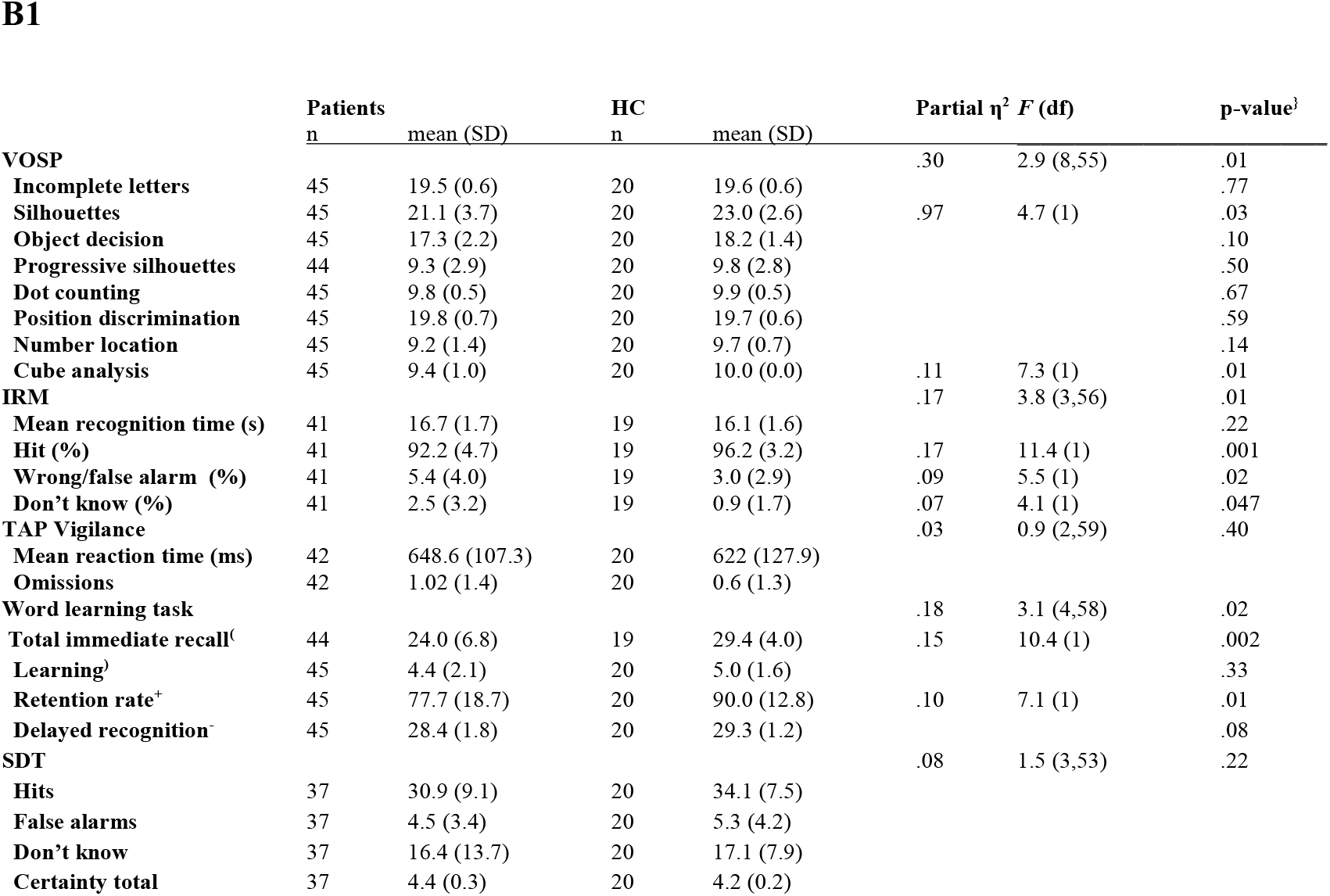
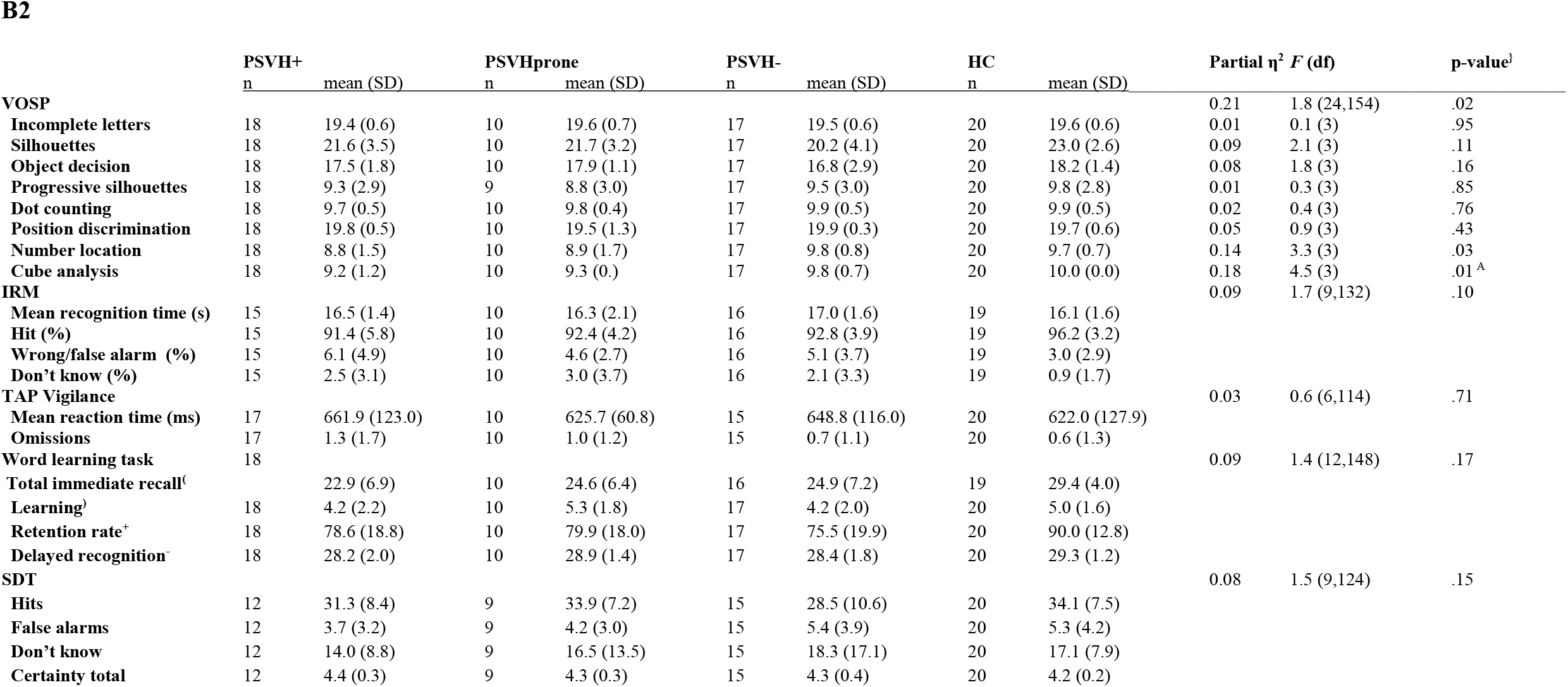
part 1: Behavioral tests.

#### Behavioral functioning

Table 1B (see end of the document) lists the mean test scores and group comparisons. The MANOVAs performed without the PSVH-prone group showed similar results (not shown). When comparing the total patient group to the HC, patients performed worse on the VOSP (*F*=2.9, p=.01); which was primarily due to their score on the Silhouettes test (*F*=4.7, p=.03) and Cube Analysis (*F*=7.3, p=.01). In addition, patients performed worse on the WLT (*F*=3.1, p=.02), having an impaired immediate recall (*F*=10.4, p=.002) and retention rate (*F*=7.1, p=.01). Patients and HC performed similarly on the TAP (*F*=0.9, p=.40) and SDT (*F*=1.5, p=.22). Finally, patients performed worse on the IRM (*F*=3.8, p=.01), having significant fewer hits (*F*=11.4, p=.001), more false alarms (*F*=5.5, p=.02), and more misses (*F*=4.1, p=.047). The recognition time did not differ between the groups.

When comparing the four groups, no significant differences were found between the patient groups for all tests. Only the VOSP showed significant results (*F*=1.8, p=.02), with the PSVH+ group performing worse than the HC on the Cube Analysis subtest (p<.05).

The number of hits on the IRM and SDT were not correlated, neither for the total group, the total patient group or each group separately (lowest p-value: .45).

### Part 2: fMRI

We included 48 participants. Of these, one HC participant was excluded due to a structural brain abnormality, one PSVH− to excessive motion. This led to 15 PSVH+, 15 PSVH− and 16 HC participants included in the further analyses. Of those, a number had also participated in part 1 (6 PSVH+, 10 PSVH− and 14 HC). Table 2A (see end of the document) depicts their demographic and illness characteristics. For the total group, the mean age was 34.6 (SD 10.0) and 70% was male. On average, patients, mainly the PSVH+, had a lower mean education level than the HC. Most patients had schizophrenia (53.3%). Of those 23.3% had a schizoaffective disorder, and 23.3% had a psychotic disorder NOS. The mean disease duration did not differ between the PSVH+ and PSVH− groups. Patients had significantly higher scores than HC for all psychotic symptoms (PANSS, QPE). PSVH+ and HC differed the most, with the scores of the PSVH− group generally in between those. Most patients used antipsychotics, mostly being one atypical antipsychotic. Binocular visual acuity ranged between 0.8 and 1.5 and contrast sensitivity between 12-16, without group differences. Two PVSH- and one HC had a visual field defect (as assessed by the HFA). These participants were not excluded, as for all, the visual field stimulated by the task was intact for at least one eye, and their binocular visual acuity and contrast sensitivity were adequate (lowest values: 1.2 and 14), and visual inspection of their movie-related activity showed acceptable occipital activity.

**Table 2:** part 2: fMRI.

**A1**
|  | **Patients**<br>**(n=30)** | **HC**<br>**(n=16)** | **Test score** | **p-value** |
| --- | --- | --- | --- | --- |
|  | **mean (SD)** | **mean (SD)** | **(df)** |  |
| Age (y) | 35.8 (9.4) | 32.4 (11.2) | 188.00 | .23 |
| Gender (n, (%)) |  |  | .34 (1) | .74 |
| Males | 20 (66.7) | 12 (75.0) |  |  |
| Females | 10 (33.3) | 4 (25.0) |  |  |
| Education* | 6.4 (1.5) | 7.5 (0.7) | 347.0 | .01 |
| MMSE<sup>#</sup> | 28.8 (1.1) | 29.7 (0.5) | 352.5 | .01 |
| Visual acuity ODS<sup>^</sup> | 1.2 (0.2) | 1.3 (0.2) | 288.0 | .24 |
| Contrast ODS<sup>~</sup> | 15.1 (1.2) | 15.4 (0.9) | 260.0 | .33 |
| Diagnosis (n, (%)) |  |  |  |  |
| Schizophrenia | 16 (53.3) |  |  |  |
| Schizoaffective disorder | 7 (23.3) |  |  |  |
| Psychosis NOS | 7 (23.3) |  |  |  |
| Disease duration (y) | 13.1 (11.0) |  |  |  |
| PANSS<sup>&</sup> |  |  | 19.1 (3,42) | .00 |
| tot pos | 16.6 (5.4) | 7.6 (0.7) | 43.1 (1) | .00 |
| tot neg | 15.0 (4.5) | 7.9 (1.6) | 35.4 (1) | .00 |
| tot gen | 35.1 (8.8) | 18.3 (1.5) | 56.6 (1) | .00 |
| tot | 66.6 (16.7) | 33.8 (2.4) | 60.1 (1) | .00 |
| QPE<sup>+</sup> |  |  | 5.3 (4,41) | .00 |
| severity VH | 6.1 (6.9) | 0 (0) | 12.4 (1) | .00 |
| severity AH | 9.6 (10.7) | 0 (0) | 12.9 (1) | .00 |
| severity all H | 16.8 (15.3) | 0 (0) | 18.9 (1) | .00 |
| severity delusions | 5.1 (5.5) | 0 (0) | 13.5 (1) | .00 |
| Use antipsychotics (n, (%)) |  |  |  |  |
| Yes | 24 (80.0) |  |  |  |
| No | 6 (20.0) |  |  |  |
| Type antipsychotics (n, (%)) |  |  |  |  |
| 1 typical | 1 (4.2) |  |  |  |
| 1 atypical | 15 (62.5) |  |  |  |
| Multiple | 8 (33.3) |  |  |  |
| Other medication<sup>l</sup> (n, (%)) |  |  | 2.39 | .30 |
| Yes, 1 | 12 (40.0) | 3 (20.0) |  |  |
| No | 11 (36.7) | 9 (60.0) |  |  |
| Multiple | 7 (23.3) | 3 (20.0) |  |  |

A2
|  | HC<br>(n=16)<br>mean (SD) | PSVH+<br>(n=15)<br>mean (SD) | PSVH-<br>(n=15)<br>mean (SD) | Test score<br>(df) | p-value |
| --- | --- | --- | --- | --- | --- |
| Age (y) | 32.4 (11.2) | 34.6 (7.9) | 37.0 (10.7) | 1.7 (2) | .43 |
| Gender (n, (%)) |  |  |  | 2.7 | .31 |
| Males | 12 (75.0) | 8 (53.3) | 12 (80.0) |  |  |
| Females | 4 (25.0) | 7 (46.7) | 3 (20.0) |  |  |
| Education* | 7.5 (0.7) | 6 (1.6) | 6.9 (1.2) | 8.9 (2) | .01 <sup>A</sup> |
| MMSE<sup>#</sup> | 29.7 (0.5) | 28.8 (1.1) | 28.9 (1.1) | 7.8 (2) | .02 |
| Visual acuity ODS<sup>^</sup> | 1.3 (0.2) | 1.1 (0.2) | 1.2 (0.2) | 4.22 (2) | .12 |
| Contrast ODS<sup>~</sup> | 15.4 (0.9) | 15.0 (1.4) | 15.2 (1.0) | 0.95 (2) | .62 |
| Diagnosis (n, (%)) |  |  |  | 1.7 | .62 |
| Schizophrenia |  | 7 (46.7) | 9 (60.0) |  |  |
| Psychosis NOS |  | 5 (33.3) | 2 (13.3) |  |  |
| Disease duration (y) |  | 11.1 (9.0) | 15.1 (12.6) | 133.0 | .41 |
| PANSS<sup>&</sup> |  |  |  | 9.8 (6,82) | .00 |
| tot pos | 7.6 (0.7) | 18.9 (4.6) | 14.3 (5.2) | 30.8 (2) | .00 <sup>A,B</sup> |
| tot neg | 7.9 (1.6) | 16.1 (5.4) | 13.9 (3.4) | 19.7 (2) | .00 <sup>A,B</sup> |
| tot gen | 18.3 (1.5) | 38.5 (8.5) | 31.7 (8.0) | 36.4 (2) | .00 <sup>A,B</sup> |
| tot | 33.8 (2.4) | 73.4 (16.1) | 59.8 (14.9) | 39.7 (2) | .00 <sup>A,B</sup> |
| QPE<sup>+</sup> |  |  |  | 21.2 (8,80) | .00 |
| severity VH | 0 (0) | 12.1 (4.3) | 0 (0) | 122.2 (2) | .00 <sup>A,D</sup> |
| severity AH | 0 (0) | 13.1 (10.8) | 6.2 (9.7) | 9.7 (2) | .00 <sup>A</sup> |
| severity all H | 0 (0) | 26.7 (12.7) | 6.8 (10.6) | 33.0 (2) | .00 <sup>A,D</sup> |
| severity delusions | 0 (0) | 5.6 (5.7) | 4.6 (5.4) | 6.9 (2) | .00 <sup>A,B</sup> |
| Use antipsychotics (n, (%)) |  |  |  |  |  |
| Yes |  | 11 (73.3) | 13 (86.7) | 0.83 (1) | .65 |
| No |  | 4 (26.7) | 2 (13.3) |  |  |
| Type antipsychotics (n, (%)) |  |  |  |  |  |
| 1 typical |  | 0 (0) | 1 (7.7) | 1.88 | .39 |
| 1 atypical |  | 6 (54.5) | 9 (69.2) |  |  |
| Multiple |  | 5 (45.5) | 3 (23.1) |  |  |
| Other medication<sup>l</sup> (n, (%)) |  |  |  | 2.78 | .63 |
| Yes, 1 | 3 (20.0) | 6 (40.0) | 6 (40.0) |  |  |
| No | 9 (60.0) | 5 (33.3) | 6 (40.0) |  |  |
| Multiple | 3 (20.0) | 4 (26.7) | 3 (20.0) |  |  |

B1
|  | **Patients**<br>**(n=30)**<br>**mean (SD)** | **HC**<br>**(n=16)**<br>**mean (SD)** | **Test score**<br>**(df)** | **p-value** |
| --- | --- | --- | --- | --- |
| **Image Recognition Movies** |  |  | 2.7 (2,43) | .08 |
| Mean recognition time (s) | 18.7 (2.5) | 18.8 (1.8) |  |  |
| % movies recognized<sup>@</sup> | 91.1 (9.0) | 94.2 (5.0) |  |  |

B2
|  | **HC**<br>**(n=16)**<br>**mean (SD)** | **PSVH+**<br>**(n=15)**<br>**mean (SD)** | **PSVH-**<br>**(n=15)**<br>**mean (SD)** | **Test score**<br>**(df)** | **p-value** |
| --- | --- | --- | --- | --- | --- |
| **Image Recognition Movies** |  |  |  | 2.0 (4,84) | .10 |
| Mean recognition time (s) | 18.8 (1.8) | 18.3 (2.6) | 19.1 (2.4) |  |  |
| % movies recognized<sup>@</sup> | 94.2 (5.0) | 91.1 (8.9) | 91.2 (9.4) |  |  |

### IRM Task performance

Directly following scanning, all participants confirmed to have performed the task according to instructions. Table 2B (see end of the document) depicts their IRM performance during fMRI scanning. Performance did not differ between groups (patients versus HC: *F*=2.7 (2,43), p=.08; three groups: *F*=2.0 (4.84), p=.10). The mean recognition time was 18.7 sec for patients, (SD 2.5), 18.8 for the HC-group (SD 1.8), 18.3 for the PSVH+ group (SD 2.6), and 19.1 for the PSVH− group (SD 2.4). The mean percentage of recognized movies was 91.1% (SD 9.0) for the patients, 94.2% (SD 5.0) for the HC-group, 91.1% (SD 8.9) for the PSVH+ group and 91.2% (SD 9.4) for the PSVH− group.

During the fMRI IRM task, three PSVH+ participants reported VH. One participant experienced seeing a dog or a donkey (amongst other things) multiple times for approximately 1 sec each time A second participant experienced seeing almost continuous light flashes, while a third participant continuously experienced a small black square moving across the entire visual field, close to the screen. Two of them also reported AH, while the third one reported a gustatory hallucination (GH; for approximately 2 min). Three additional PSVH+ participants reported non-visual hallucinations: one of them reported both AH (once during a period of a few seconds) and tactile hallucinations (TH; twice during a period of a few seconds), one reported AH (almost continuously), while a third one reported TH (once during a period of a few seconds).

Three of the PSVH− participants reported non-visual hallucinations: one experienced AH ((almost) continuously) and TH (3-4 times for a couple of seconds), one experienced AH (hearing twice two words), and one participant experienced TH (once per movie during a period of a few seconds). As they reported that they were able to perform the task adequately despite their hallucinations, they were not excluded.

#### Brain activity during movie presentation and during the period of recognition

Whole brain analysis revealed increased activation in all groups during the entire period of the movies, and the period of recognition (i.e. from pop-out until movie ending). Details on these analyses are given in Suppl. 2A,B and 1C,D, respectively. Here, we focus on the activity around pop-out using a whole brain and ROI-based FIR analysis.

##### Whole brain FIR analysis

Fig. 2 depicts the mean group whole brain activity around pop-out. HC and PSVH− showed a highly similar pattern, starting with fusiform gyrus (FG) activation from −3 and −2 sec before pop-out, with subsequent more widespread occipital, temporal, parietal and frontal activation. In contrast, the PSVH+ group did not show significant FG activation until pop-out. Moreover, subsequent activity in other areas was not as pronounced.

**Fig. 2:**
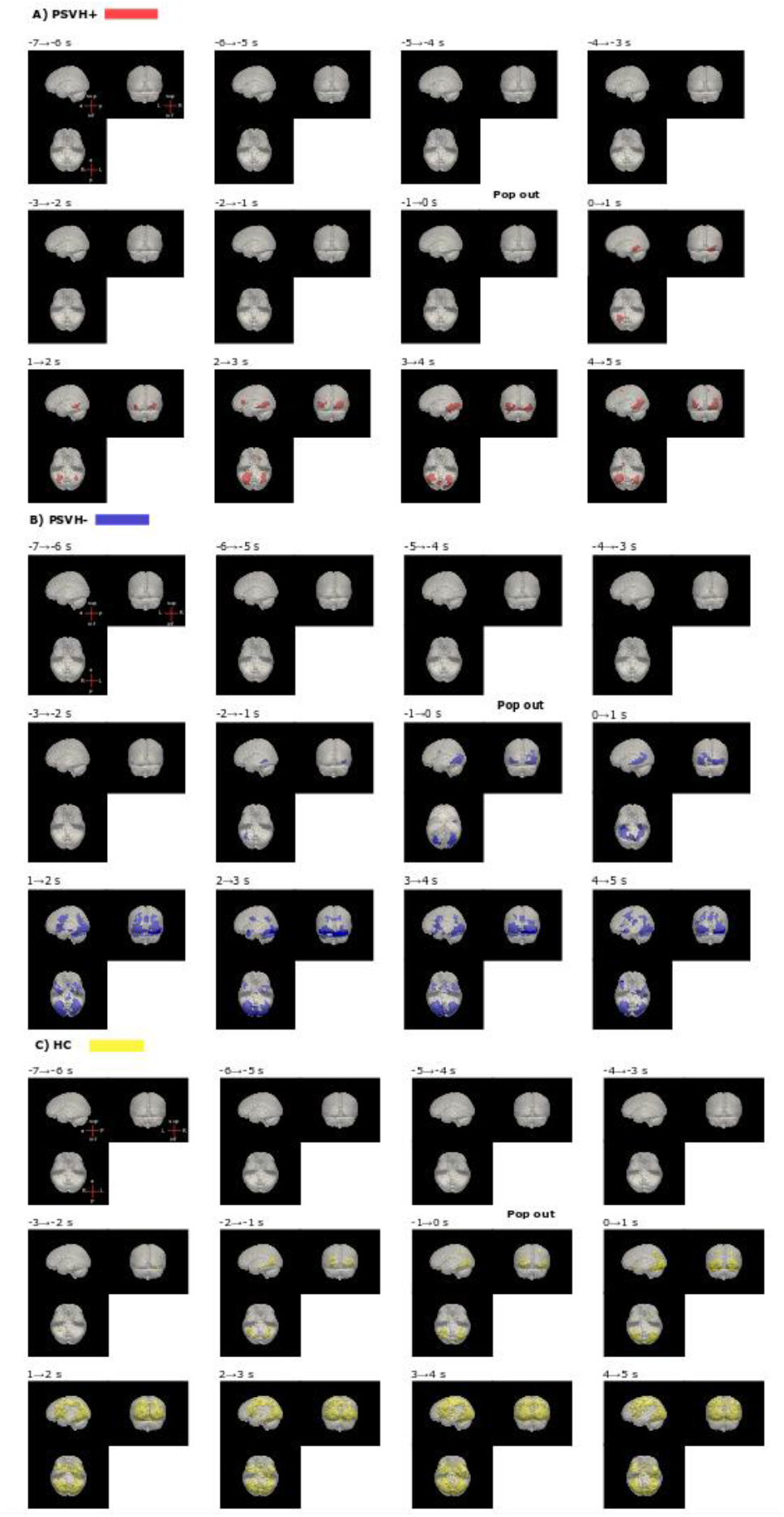
fMRI whole brain activity patterns around pop out. Legend: Image Recognition Movies task: whole brain fMRI activity patterns (Finite Impulse Response analysis) per group around pop-out, from 7 s before pop-out until 5 s after pop-out; 1 s per time bin. Per time bin: upper image: view from the left and posterior; image below: view from below. A) PSVH+: patients with psychosis with visual hallucinations, B) PSVH−: patients with psychosis without visual hallucinations, C) HC: healthy controls. All analyses: p<0.001 on voxel level and 0.05 FWE cluster-wise correction.

##### ROI-based FIR analysis

Fig. 3 displays the group comparisons of the ROI time series. In general, activity started to increase several seconds before pop-out, followed by a subsequent decrease after pop-out. This was the case in all groups and in nearly all ROIs. However, a markedly different pattern of activity was present in the angular gyri, and to a lesser extent in the supramarginal gyri post. Activity decreased just before pop-out, followed by an increase after pop-out.

**Fig. 3:**
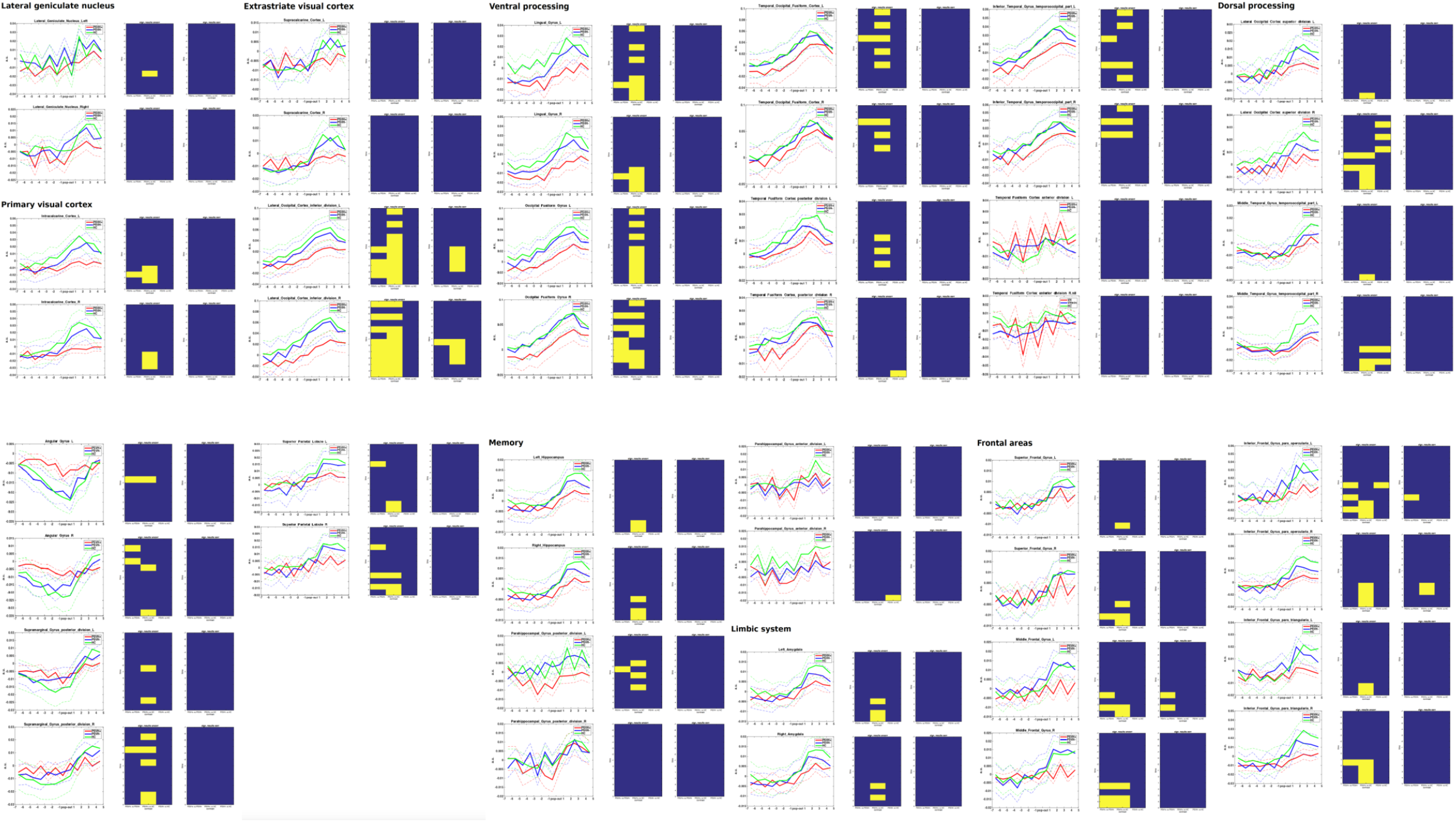
ROI timeseries around pop out including group comparisons. Legend: Image Recognition Movies task: activity patterns around pop-out for predefined regions of interest (ROI’s) that play a role in visual object recognition, including the lateral geniculate nucleus (LGN, thalamus), V1, ventral and dorsal processing, memory and limbic-related areas, and frontal areas. PSVH+: patients with psychosis with visual hallucinations, PSVH−: patients with psychosis without visual hallucinations, HC=healthy controls. Per ROI: left: mean group activation levels (in arbitrary units, corrected for ‘baseline’ activity of the whole movie) per group per time bin (from 7 s before pop-out until 5 sec after pop-out; 1 sec per time bin). Middle: group comparisons: PSVH+ vs PSVH−, PSVH+ vs HC, PSVH− vs HC, uncorrected. Right: group comparisons, corrected for number of time bins and contrasts using FWE correction. All comparisons: two-tailed, p<0.05

In many ROIs, across all parts of the visual system, activity for the PSVH+ group tended to increase less than the PSVH− and HC-group both before and after pop-out. In the angular gyri (and to a lesser extent the supramarginal gyri post), activity in the PSVH+ group tended to decrease less before pop-out, and also increase less after pop-out.

Examining the differences between groups, the inferior LOC activity increased less for the PSVH+ group compared to the other two groups just before pop-out (1 sec before pop-out until pop-out). Compared to the PSVH− group, this includes the right inferior LOC. Compared to the HC-group, this includes both the left and right inferior LOC. Furthermore, from pop-out until 3 sec after pop-out, activity in the bilateral inferior LOC increased less in the PSVH+ group than in the HC-group. After pop-out, bilateral middle frontal gyri activity increased less for the PSVH+ group than in PSVH−, and the bilateral opercular part of the inferior frontal gyrus increased less compared to both the PSVH− group and the HC-group.

In short, the PSVH+ group showed less responsiveness to the stimulus popping out of noise.

## Data and code availability

The raw data and code belonging to this study are available from the corresponding author upon reasonable request.

## Discussion

In accordance with our hypothesis, our study first showed that psychosis patients generally have worse behavioral performance on tests assessing visual perception, attention or working memory. Second, psychosis patients with a predisposition for VH and patients without VH had the same behavioral performance. Third, prior to image recognition, patients with a predisposition for VH – when compared to those without VH – showed reduced responsiveness of the occipital areas, which was most prominent in the object recognition region LOC. However, we found no evidence for an accompanying differential prefrontal activation. The absence of a behavioral deficit in patients with a predisposition for VH might be explained by a trend towards reduced suppression of the angular gyri around image recognition in this group. This finding may indicate a compensatory mechanism that involves these visual memory-related regions. Below, a more detailed discussion is provided.

### Reduced occipital responsiveness in PSVH+ around pop-out

In all groups, during the movies and during the period of recognition, the ventral visual areas showed robust task-related activation (Suppl. Figs. 1C,D and 2A,B). Moreover, the ROI-based FIR analyses depict that generally, visual areas showed increasing activity before pop-out (Fig. 3). Critically, the subsequent decrease after pop-out indicates that the increase in activity was not just caused by increasing object contrast.

Importantly, and in line with our hypothesis, inferior LOC activity increased less in the PSVH+ group around pop-out. The LOC plays a central role in object recognition ^34^ and receives input via the ventral stream and, via feedback projections from the prefrontal cortex, from the dorsal stream ^35^. Thus, our results indicate reduced occipital responsiveness in the PSVH+ group, which reached its lowest level in the LOC.

Theoretically, the reduced occipital activation could have been caused by a ceiling effect. Also hallucinations during scanning may have reduced activity patterns caused by the task ^36^. In our study, only three PSVH+ participants experienced VH during scanning. Repeating the FIR ROI analyses while excluding them gave very similar results (Suppl. 3), indicating that the VH experienced during scanning are not responsible for the reduced occipital responsiveness in the PSVH+ group. Previous fMRI, event-related potential and (ERP) and magnetoencephalographical studies also found reduced task-related occipital activity in schizophrenia ^9,35,37,38^. Because the PSVH− group’s results did not differ from those of the HC, our study goes beyond these findings by suggesting that the reduced occipital responsiveness may specifically underlie the predisposition for VH, rather than to psychosis in general. Patients with schizophrenia had lower V2-V4 activation than controls when visually integrating a contour consisting of individual elements ^9^. Moreover, patients showed reduced ERP generation over the LOC when recognizing complete objects based on fragments ^35,37^. Path analyses in a combined ERP-fMRI study showed that schizophrenia had a significant effect on the dorsal stream and the frontal cortex, but not directly on the LOC ^35^. This suggests that the input to the LOC may be impaired, rather than the LOC itself. This is in line with our present finding, where the reduced LOC activity might primarily reflect the early visual processing deficit that we also observed (but that did not reach statistical significance). An explanation for why the LOC activity was significantly reduced while that of pre-LOC visual areas was not, may be found in the more specific role of the LOC in object recognition. Moreover, the LOC has a larger cortical magnification factor than earlier visual areas, implying that the object movies activated a relatively large part of the LOC. Moreover, due to its large receptive fields ^39^, it may have been less activated by the dynamic noise in the movies, reducing response variance.

### Possible biological substrates for the reduced occipital responsiveness

Both neurotransmitters and anatomical alterations may contribute to the reduced occipital responsiveness. First, patients with schizophrenia show decreased GABA concentrations in the visual cortex ^40^, which relate to impaired surround suppression. In the case of our movies, suppression will serve to single out foreground (i.e. relevant object-related signal) from background. Interestingly, it has been proposed that decreased GABA concentrations are downstream effects of impaired NMDA receptor neurotransmission ^40,41^, which is involved in the pathophysiology of schizophrenia and relates to the occurrence of hallucinations ^42^. Moreover, dopaminergic dysregulation in both cortical and subcortical systems plays an important role in schizophrenia ^43^, with a predisposition to hallucinations mostly being related to increased striatal dopamine transmission ^44^. Dopamine improves the occipital signal-to-noise-ratio and the efficacy of visual decision making ^45,46^. Lastly, patients with schizophrenia have smaller population receptive field sizes in visual areas ^47^, increased functional segregation in the occipital cortex ^48,49^ and thinner cortices of early visual areas and the LOC ^50^ than controls. However, it is not known whether these various anatomical alterations, possibly in a more extreme form, would predispose patients to experience VH.

### VH are not specifically related to behavioral impaired visual perception, possibly due to a compensatory memory-related mechanism

Part 1 confirmed earlier studies by showing that patients had worse behavioral visual object and space perception and working memory than HC ^7–12,14,51^. However, test scores did not differ between the PSVH+ and PSVH− groups, implying that a predisposition for VH is not necessarily related to impaired visual perception, attention and working memory. However, we did find reduced occipital activation during image recognition in PSVH+. An explanation why this did not lead to impaired behavioral visual perception may be found in the activity patterns of the angular gyri around pop-out. This region belongs to the dorsal pathway and may also contribute to object recognition ^52,53^. In the HC-group, angular gyri activity decreased markedly around pop-out, indicating that the angular gyri are normally suppressed around pop-out. In this, the PSVH− group resembled the HC, while in the PSVH+ group activity was much less suppressed. As the angular gyri, being part of the Default Mode Network (DMN), are involved with visual memory retrieval, this may indicate a heightened reliance on visual memory-related information ^54,55^. It might also indicate reduced suppression of salient features outside the image to be recognized ^56^. However, the latter would likely further impair visual processing, which is not supported by our behavioral results. Therefore, compensatory reliance on memory might have led to the non-deviant behavioral visual perception in the PSVH+ group when compared to PSVH− group.

### Visual processing deficits predispose to VH in psychosis

Various models state that impaired visual processing contributes to experiencing complex VH. For example, the ‘Perception and Attention Deficit’ model states that to choose the correct object, amongst multiple potentially observed objects, adequate object perception and attentional object-binding are necessary (12). Co-occurring impairments may lead to choosing an erroneous object, resulting in VH. Diederich et al considered VH as a dysregulation of the gating and filtering of external and internal percepts ^57^.

Impaired vision, reduced activation of V1 and aberrant activation of higher visual areas would contribute to this dysregulation. A third model postulates that dysfunctioning of the visual network and vision-related networks may lead to an erroneous interpretation of ambiguous signals, and thus to VH ^58^.

A relatively recent theory for explaining VH in psychosis is based on Bayesian inference and predictive coding (PC). This notion states that visual perceptions come about by integrating predictions (based on prior beliefs and presumably generated by the prefrontal cortex) with incoming sensory input coming from visual areas ^59^. A mismatch between prediction and visual input results in an error signal that is used to update one’s beliefs about the world.

A relative imbalance between predictions and visual input may bias one’s inferences towards one’s predictions, resulting in VH. While our study was not designed to test this theory we will consider our results in its terms. The reduced occipital signals may indicate a reduced precision of the visual input, which could result in the imbalance that causes VH. The reduced angular gyri suppression could indicate a compensatory reliance on priors. However, the reduced occipital signals may also indicate a diminished error signal, due to a reduced prior that can match a broad array of (normal) incoming visual input. Indeed, both reduced and strong priors have been suggested in psychotic VH generation ^59–61^. A final theory we want to consider is that of a change in contextual modulation (CM) in schizophrenia ^62^. CM is defined as influences that modulate the sensitivity of a cell to its normal input, but that do not normally lead to an action potential in the cell itself. Our results fit this theory quite well, as a CM may result in a reduced overall gain that can explain both the reduced occipital increase in activity and the reduced angular gyri decrease in activity around pop-out. Notably, CM may also affect the precision of neural signals ^62^. Since the visual input and priors of PC theory are both neural signals, explanations in terms of CM and PC need not be mutually exclusive. Concluding, this topic warrants further research with specifically designed experiments.

### Comparing visual processing in patients with VH between psychosis and PD

Our hypotheses were based on observations in PD (16). Similar to the PD patients with VH, our present PSVH+ group had reduced LOC activation around pop-out. However, PD patients with VH also had reduced frontal activation and behavior-impaired visual perception and attention ^16–18^; for which we found no evidence in PSVH+ participants. During a behavioral task, PD patients with VH showed reduced sensory evidence accumulation ^63^, which could be interpreted as a reduced sensory precision. Interestingly, both the impairments in perception and attention in PD relate to organic pathology. VH in PD are associated with baseline occipitotemporoparietal hypometabolism ^64^. VH severity relates to temporal and parietal Lewy Body depositions ^65–67^, corresponding with visuospatial and visuoperceptive deficits. Patients with PD dementia, being more at risk for VH, have higher occipital atrophy ^68,69^. Furthermore, VH in PD closely correlates with attentional deficits, which relate to reduced acetylcholine levels ^70–72^. Moreover, dopaminergic medication may induce or exacerbate VH ^73^. Interestingly, VH in PD are also related to dysfunction of the visual system and DMN ^74^. In a visual task that evokes misperceptions, considered a surrogate for VH, PD patients with VH showed higher DMN activity and hyper-connectivity between the DMN and visual system. Thus, VH in PD are related to anatomical alterations, altered neurotransmitters and dysfunctional vision-related networks. Remarkably, VH in PD are associated with impaired behavioral vision and attention, whereas such impairments are associated with schizophrenia in general, but not specifically with VH. This suggests that even though both PD and schizophrenia show impaired visual processing that predisposes for VH, there are subtle differences in their underlying mechanisms.

### Limitations and future studies

Our study has some limitations. First, the precise influence of antipsychotics is hard to quantify: antipsychotics antagonize dopamine receptors, while, as stated above, dopamine improves the occipital signal-to-noise-ratio and the efficacy of visual decision making ^45,46^. Second, the IRM task did not assess eye movements, whereas differential eye movements may result in differential occipital responsiveness. However, performance on the central fixation task did not differ between the patient groups, suggesting no differences in eye movements.

Given the differential fMRI activity between the PSVH+ and PSVH− groups, we recommend future studies to take VH into account when investigating schizophrenia. Furthermore, predictive coding, functional network and structural approaches may contribute to further our understanding the role of abnormal visual processing in patients with schizophrenia. While our present image-in-noise recognition task is attractive because of its simplicity for the participant, it may be possible to further improve it by employing a forced-choice recognition task, varying properties of the noise ^75^, and using further image and object perturbations ^76,77^ to more precisely pin-point the processing impairment(s) associated with VH.

## Conclusion

Our study is the first to show that a predisposition for VH in psychosis is associated with impaired visual processing, most notably in the object recognition region LOC. Remarkably, these abnormalities were found in the absence of behavior-impaired visual perception, attention or working memory. Unlike in PD, we found no evidence for accompanying differential prefrontal activation around image recognition. However, the angular gyri showed a trend for reduced suppression. This may suggest a compensatory mechanism from these visual memory-related regions, explaining the non-deviant behavioral visual perception. The impaired visual processing may contribute to erroneous visual perceptions. The higher reliance on memory may predispose patients towards errors in assigning origin (internal instead of external) to perceptions. Their combination may contribute to VH in psychosis.

## Supporting information

STROBE

## Acknowledgements

We would like to thank Wendy Groot Jebbink, Martijn Majoor, Wouter Staal, and Anita Sibeijn-Kuiper for their help in fMRI data acquisition.

## Supplementary material

