## Supplementary material for "Impaired visual processing in psychosis patients with a predisposition for visual hallucinations"

**Content**

Suppl. 1: Image Recognition Movies task: activity during recognition
Suppl. 2: Image Recognition Movies task: activity during the movies
Suppl. 3: FIR ROI analysis for the left and right LOC inferior, without the 3 PSVH+ participants who experienced VH during scanning

#### Suppl. 1: Image Recognition Movies task: activity during recognition

**A: IRM paradigm** (see manuscript for description)

**B: model for recognition**

First, per participant a General Linear Model (GLM) was built, with five conditions entered in the following order: 1) pop out, 2) recognition (pop out until movie end), 3) movie (for overall task-related activity), 4) control movie, 5) static noise at the beginning of each run. Per subject, the contrast 'recognition' versus baseline (red fixation cross on a black screen) was obtained. Next, group means were determined (Suppl. 1 C1). Subsequently, ANOVA compared groups with $p<0.001$ on voxel level and 0.05 FWE cluster-wise correction (Suppl. 1C2).

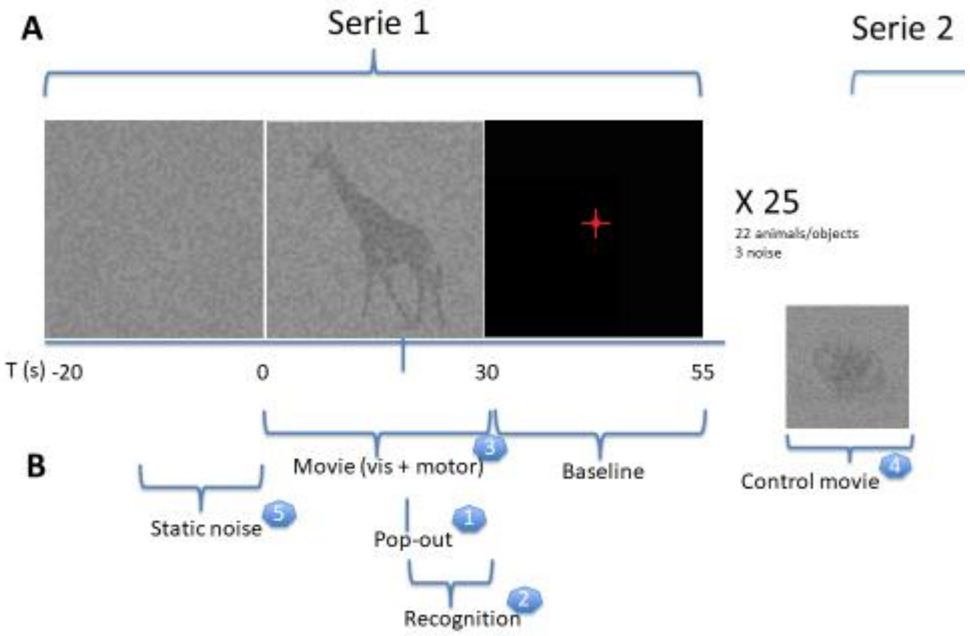


Recognition: recognition (2) versus baseline

### C: Activity during recognition

#### B1: mean group activity

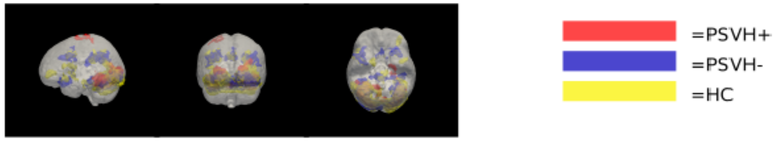


#### B2: group comparisons

**PSVH+ vs PSVH-**

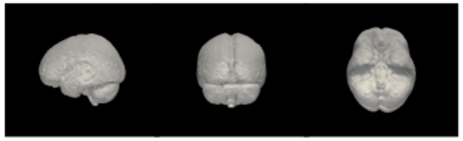

**PSVH+ vs HC**

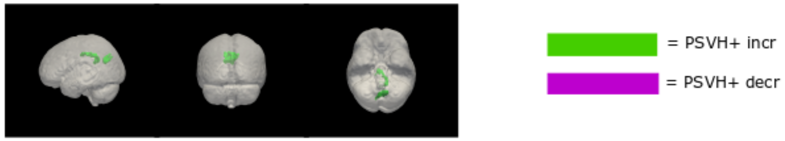


**PSVH- vs HC**

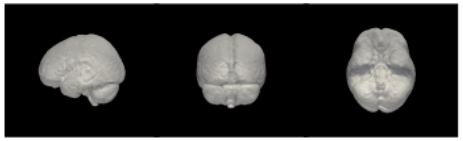

### D1: per group

| Group | Anatomical region | MNI-coord | Voxels (n) | Peak T (voxel) | p-value (FWE cluster corr) |
|---|---|---|---|---|---|
| **PSVH+** | Lingual gyrus l | -26, -53, -8 | 200 | 7.52 | 0.000 |
| | Lingual gyrus l | -14, -53, -5 | | 5.46 | |
| | Lingual gyrus l | -14, -68, -5 | | 4.98 | |
| | Sup temp sulc post r | 58, -41, 7 | 277 | 6.32 | 0.000 |
| | Sup temp sulc post r | 55, -23, -5 | | 6.14 | |
| | Sup temp gyr post r | 67, -32, 4 | | 5.47 | |
| | Lingual gyrus r | 13, -44, -8 | 211 | 6.18 | 0.000 |
| | Lingual gyrus r | 22, -65, -5 | | 5.85 | |
| | Cerebellum r I-IV | 4, -47, -2 | | 4.50 | |
| **PSVH-** | Sup temp sulc post l | -62, -41, 4 | 288 | 8.59 | 0.000 |
| | Sup temp gyr post l | -53, -23, -2 | | 5.63 | |
| | Angular gyrus l | -47, -50, 13 | | 5.47 | |
| | Lingual gyrus l/intracalc cortex | -17, -71, 1 | 269 | 6.41 | 0.000 |
| | Lingual gyrus r | 13, -71, -2 | | 5.53 | |
| | Lingual gyrus l | -20, -47, -2 | | 3.98 | |
| | Inf front gyr, operc r | 58, 22, 13 | 120 | 5.95 | 0.001 |
| | Frontal pole lat r | 46, 46, -2 | | 5.76 | |
| | Inf front gyr, triang r | 49, 28, -2 | | 4.90 | |
| | Mid temp gyr post r | 64, -32, -2 | 135 | 5.86 | 0.001 |
| | Sup temp gyr post r | 46, -20, -5 | | 5.54 | |
| | Sup temp sulc mid r | 64, -14, -8 | | 5.41 | |
| **HC** | Mid temp gyr post l | -56, -35, -5 | 232 | 7.44 | 0.000 |
| | Sup temp sulc mid l | -47, -17, -11 | | 4.54 | |
| | Sup temp gyrus post WM l | -47, -38, 7 | | 4.17 | |
| | Mid temp gyr post r | 46, -32, -5 | 581 | 7.34 | 0.000 |
| | Inferior parietal lobe r | 55, -44, 28 | | 6.61 | |
| | Mid temp gyr t-o r | 55, -41, 4 | | 5.13 | |
| | Cuneus WM r | 22, -68, 16 | 933 | 6.10 | 0.000 |
| | Lingual gyrus l | -14, -59, -8 | | 5.86 | |
| | Cuneal cortex r | 10, -77, 25 | | 5.67 | |
| | Supramarginal gyrus post l | -47, -47, 28 | 140 | 4.55 | 0.002 |
| | Inferior parietal lobe l | -56, -44, 37 | | 4.49 | |
| | Angular gyrus l | -56, -50, 16 | | 4.48 | |

### D2: group comparisons

| Comparison | Increased activation<br>Anatomical region | MNI-coord | Voxels<br>(n) | Peak T<br>(voxel) | p-value<br>(FWE cluster corr) | Decreased activation<br>Anatomical region | MNI-coord | Voxels<br>(n) | Peak T<br>(voxel) | p-value<br>(FWE cluster corr) |
|---|---|---|---|---|---|---|---|---|---|---|
| **PSVH+ vs PSVH-** | - | | | | | - | | | | |
| **PSVH+ vs HC** | Lingual gyrus l | -11, -80, -8 | 83 | 4.17 | 0.045 | Inferior parietal lobe r | 55, -44, 28 | 83 | 4.76 | 0.045 |
| | Lingual gyrus r | 7, -80, -8 | | 3.65 | | | | | | |
| **PSVH- vs HC** | - | | | | | - | | | | |

**Suppl. 2: Image Recognition Movies task: activity during the movies**

**A: Activity during the movies**

**A1: mean group activity**

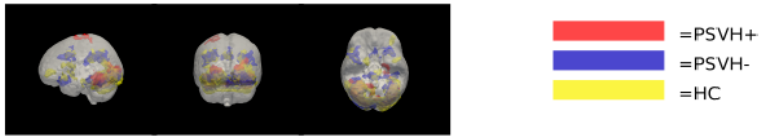


**A2: group comparisons**

**PSVH+ vs PSVH-**

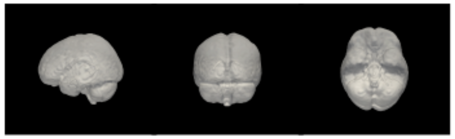

**PSVH+ vs HC**

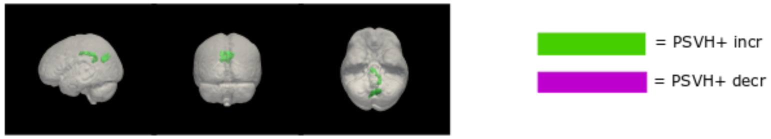


**PSVH- vs HC**

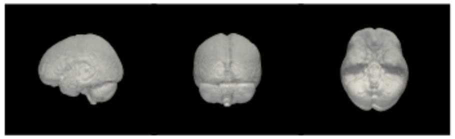

### B: Activated regions during the movies

### B1: per group

| Group | Anatomical region | MNI-coord | Voxels (n) | Peak T (voxel level) | p-value (FWE cluster corr) |
|---|---|---|---|---|---|
| **PSVH+** | Cerebellum VI r | 31, -47, -26 | 1302 | 8.70 | 0.000 |
| | Occipital fusiform gyrus r | 40, -65, -14 | | 7.97 | |
| | Cerebellum I-IV r | 7, -53, -17 | | 7.85 | |
| | Cerebellum crus I l | -35, -71, -23 | 853 | 8.33 | 0.000 |
| | Temporo-occipital fusiform cort l | -35, -53, -17 | | 7.62 | |
| | Occipital fusiform gyrus l | -20, -89, -11 | | 6.48 | |
| | Premotor cortex l | -20, -5, 76 | 70 | 6.91 | 0.016 |
| | Central sulcus l | -20, -26, 79 | | 5.69 | |
| | Precentral gyrus l | -29, -11, 70 | | 5.48 | |
| **PSVH-** | LOC inf l | -38, -71, -5 | 2358 | 17.11 | 0.000 |
| | Occipital fusiform gyrus l | -29, -80, -17 | | 12.40 | |
| | Occipital fusiform gyrus r | 19, -86, -11 | | 12.28 | |
| | Superior colliculus r | 7, -26, -8 | 160 | 7.57 | 0.000 |
| | Thalamus l WM | -23, -29, -2 | | 6.46 | |
| | Superior colliculus l | -5, -32, -5 | | 5.79 | |
| | Superior parietal lobe r | 28, -47, 46 | 293 | 6.61 | 0.000 |
| | Inferior parietal lobe r | 46, -41, 46 | | 5.21 | |
| | Inferior parietal lobe r | 22, -56, 37 | | 5.09 | |
| | Superior parietal lobe l | -32, -41, 43 | 381 | 6.61 | 0.000 |
| | Superior parietal lobe l | -26, -50, 46 | | 5.77 | |
| | Inferior parietal lobe l | -44, -44, 43 | | 5.64 | |
| | Temporal pole/Frontal orbital cort/Insula r | 43, 16, -14 | 142 | 6.56 | 0.000 |
| | Insular cortex ant r | 37, 19, 1 | | 5.22 | |
| | Inf front gyr, triang r | 34, 34, 4 | | 4.72 | |
| | Superior frontal gyrus post l | -5, 13, 55 | 266 | 6.55 | 0.000 |
| | Medial frontal gyrus r | 10, 13, 49 | | 5.59 | |
| | Medial frontal gyrus l | -2, 28, 49 | | 5.45 | |
| | Frontoparietal operculum r | 43, 4, 7 | 213 | 6.06 | 0.000 |
| | Precentral sulcus r | 40, 7, 31 | | 5.25 | |
| | Precentral sulcus r | 37, 1, 43 | | 5.18 | |
| | WM putamen/insular cortex ant l | -23, 22, -8 | 124 | 5.74 | 0.001 |
| | Frontal orbital cortex l | -35, 25, -8 | | 5.37 | |
| **HC** | Occipital fusiform gyrus l | -32, -77, -17 | 4150 | 14.03 | 0.000 |
| | Lingual gyrus l | -11, -86, -11 | | 13.13 | |
| | Occipital fusiform gyrus l | -23, -86, -11 | | 13.11 | |
| | Superior colliculus r | 4, -29, -8 | 109 | 7.31 | 0.002 |

| | | | | |
|---|---|---|---|---|
| WM thalamus l | -11, -20, -5 | | 5.40 | |
| Cerebellum I-IV l | -2, -41, -14 | | 4.66 | |
| Frontal pole l | -38, 46, 16 | 78 | 6.20 | 0.009 |
| Inferior parietal lobe l | -26, -41, 40 | 210 | 5.59 | 0.000 |
| Inferior parietal lobe l | -44, -32, 40 | | 5.48 | |
| Superior parietal lobe l | -23, -62, 58 | | 5.35 | |
| Middle frontal gyrus lat r | 52, 25, 34 | 229 | 5.48 | 0.000 |
| Precentral gyrus r | 46, 4, 28 | | 4.73 | |
| Inferior frontal gyrus, operc r | 43, 4, 19 | | 4.31 | |
| Medial frontal gyrus r | 7, 10, 58 | 168 | 5.42 | 0.000 |
| SMA l | -5, 1, 55 | | 5.21 | |
| Medial frontal gyrus l | -8, 16, 46 | | 4.33 | |
| Caudate nucleus r | 13, 4, 13 | 61 | 5.42 | 0.026 |
| WM putamen r | 19, 7, 7 | | 4.76 | |
| Pallidum r | 16, -5, -2 | | 4.50 | |
| Insular cortex ant l | -32, 22, 4 | 72 | 5.27 | 0.013 |
| Inf front gyr, triang l | -38, 22, 13 | | 4.58 | |
| Precentral sulcus l | -47, 4, 28 | 57 | 5.19 | 0.034 |

### B2: group comparisons

| **Comparison** | **Increased activation** | | | | | **Decreased activation** | | | | | |
|---|---|---|---|---|---|---|---|---|---|---|---|
| | **Anatomical region** | **MNI-coord** | **Voxels (n)** | **Peak T (voxel)** | **p-value (FWE cluster corr)** | **Anatomical region** | **MNI-coord** | **Voxels** | **Peak T (n)** | **p-value (voxel)** | **(FWE cluster corr)** |
| **PSVH+ vs PSVH-** | - | | | | | - | | | | | |
| **PSVH+ vs HC** | Cingulate cortex post r | 7, -17, 40 | 160 | 4.07 | 0.001 | - | | | | | |
| | Cingulate cortex post r | 1, -47, 25 | | 4.06 | | | | | | | |
| | Cingulate cortex post l | -14, -44, 37 | | 3.83 | | | | | | | |
| | Precuneous cortex l | -8, -65, 25 | 126 | 3.98 | 0.003 | | | | | | |
| | Parietooccipital fissure r | 4, -71, 28 | | 3.83 | | | | | | | |
| | Cuneal cortex | 13, -80, 34 | | 3.44 | | | | | | | |
| **PSVH- vs HC** | - | | | | | - | | | | | |

**Suppl. 3: FIR ROI analysis for the left and right LOC inferior, without the 3 PSVH+ participants who experienced VH during scanning**

**A: left**

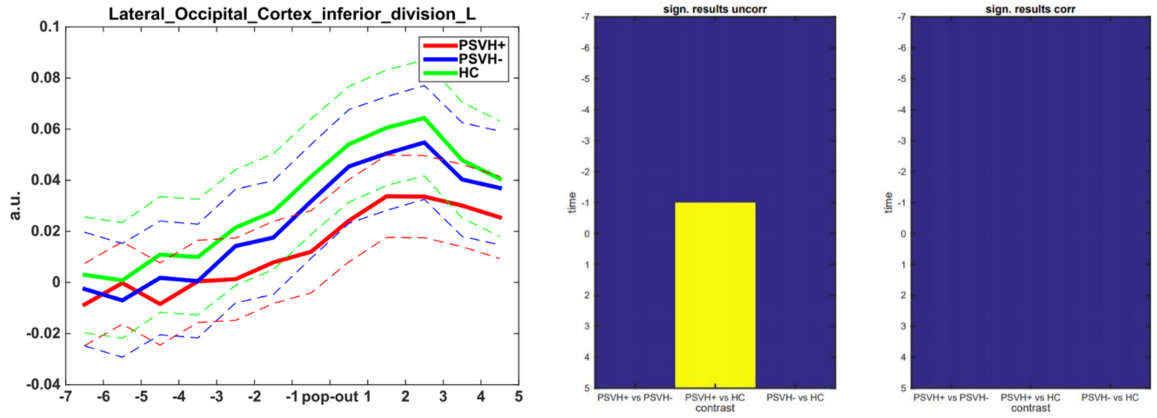


**B: right**

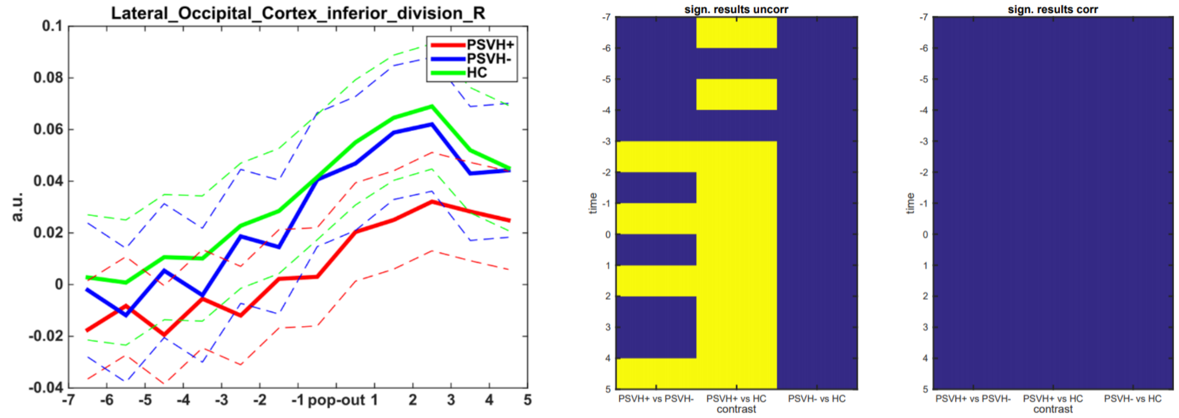
